# Discovering Signature Disease Trajectories in Pancreatic Cancer and Soft-tissue Sarcoma from Longitudinal Patient Records

**DOI:** 10.1101/2025.02.19.25322573

**Authors:** Liwei Wang, Rui Li, Andrew Wen, Qiuhao Lu, Jinlian Wang, Xiaoyang Ruan, Adriana Gamboa, Neha Malik, Christina L. Roland, Matthew H.G. Katz, Heather Lyu, Hongfang Liu

**Affiliations:** McWilliams School of Biomedical Informatics, The University of Texas Health Science Center at Houston, Houston, TX, USA; Department of Surgical Oncology, Division of Surgery, The University of Texas MD Anderson Cancer Center, Houston, TX, USA

## Abstract

Understanding the disease trajectories of specific diseases can provide important clinical insights. In this paper, we aimed to discover signature disease trajectories of 3 rare cancer types: pancreatic cancer, soft tissue sarcoma (STS) of the trunk and extremity (STS-TE), and STS of the abdomen and retroperitoneum (STS-AR), leveraging IQVIA Oncology Electronic Medical Record. We identified significant diagnosis pairs in patients with these cancers through matched cohort sampling, statistical computation, right-tailed binomial hypothesis test, and visualized trajectories up to 3 progressions. Results included 266 significant diagnosis pairs for pancreatic cancer, 130 for STS-TE, and 118 for STS-AR. We further found 44 2-hop (i.e., 2- progression) and 136 3-hop trajectories before pancreatic cancer, 36 2-hop and 37 3-hop trajectories before STS-TE, and 17 2-hop and 5 3-hop trajectories before STS-AR. Meanwhile, we found 54 2-hop and 129 3-hop trajectories following pancreatic cancer, 11 2-hop and 17 3- hop trajectories following STS-TE, 5 2-hop and 0 3-hop trajectories following STS-AR. Systematic validation of discovered trajectories with the UTHealth Electronic Health Records confirmed the feasibility and reliability of our method. Our result suggested that some key clinical features can potentially serve as early markers of rare cancers. This approach is generalizable to other disease types and real-world longitudinal patient records.

## INTRODUCTION

Rare cancers are defined as those with an incidence of fewer than 15 cases per 100,000 people per year.^1, 2^ Pancreatic cancer and soft-tissue sarcoma (STS) are rare cancers that have an incidence of 8 cases per 100,000 people and 1.8 to 3 cases per 100,000 people, respectively.^3–5^ Most clinicians have limited experience with these rare diseases, making diagnosis and treatment challenging. Large real-world data sources, such as electronic health records (EHRs), provide a massive amount of information that can potentially be leveraged to determine the patterns of diagnoses and treatments for rare tumors that can serve as clinical decision aids.

The study of large, diverse population-based datasets can help uncover disease trajectories.^6, 7^ In one study, large-scale longitudinal registry data were used to identify 1,171 unique condensed disease trajectories that were grouped into 10 major clusters that represented the most common pathways for disease progression.^8^ This approach has also been applied in the cancer domain; researchers have used a large patient registry to map the significant associations between diseases occurring before cancer diagnoses and created longitudinal disease trajectories that captured 168 temporal diagnosis patterns across 7 cancer types.^9^ Additionally, an analysis of disease progression across more than 300 conditions in a dataset of 10.4 million inpatients uncovered an unexpected association between schizophrenia and rhabdomyolysis, highlighting the ability of these large-scale methods to reveal novel disease correlations.^10^ The methods can also evaluate interconnected disease pathways; a recent study mapped temporal correlations of diseases commonly leading to hospital deaths.^11^ These broad, overarching patterns of disease progression found in studies analyzing large-scale longitudinal records can help inform both research and clinical practice.^8–12^

Understanding general disease trajectories suggests the importance of identifying specific disease trajectories, particularly for rare diseases. Disease-specific trajectories can reveal subtle temporal patterns that can help clinicians refine diagnosis, treatment, and prognosis for a specific condition. These trajectories can also support the development of predictive models and algorithms to improve early detection and aid clinical decision-making. Nevertheless, studies on disease-specific trajectories remain limited. A recent study introduced a tool for visualizing and analyzing bladder cancer trajectories.^13^ However, the selected study cohort with only bladder cancer patients allowed for commonly cooccurring medical problems to be filtered out, compromising the discovery of complete disease progression patterns. For example, hematuria, the most typical symptom of bladder cancer, was excluded from the final disease trajectory, which may not fully capture the clinical path leading to a bladder cancer diagnosis.^14,15^

To the best of our knowledge, there are no published studies on the signature disease trajectories of other rare cancers, including pancreatic cancer and STS. The aim of this study was to develop an approach to identify and describe the signature disease trajectories for pancreatic cancer, STS of the trunk and extremity (STS-TE), and STS of the abdomen and retroperitoneum (STS-AR) by leveraging large-scale EHR data.

## METHODS

### Data sources

IQVIA data were used to determine and visualize disease trajectories, and the UTHealth Observational Medical Outcomes Partnership (OMOP) common data model (CDM) dataset was used for trajectory validation. This study was approved by the Institutional Review Board of UTHealth.

### IQVIA Oncology Electronic Medical Record

Data in the IQVIA Oncology Electronic Medical Record (EMR) dataset are sourced from oncology-specific EMR vendors at 3 types of sites: large practice, site services partner, and multiple practices/locations. Data from all sources are provided in a structured format and mapped to a common data model (CDM). These data include clinically rich information across various cancer types. The dataset includes clinical information from over 1.2 million patients from 2015 through 2023.

### UTHealth OMOP CDM

EHR data from 2005 to 2024 from the McGovern Medical School at The University of Texas Health Sciences Center was transformed using the Observational Health Data Sciences and Informatics’ OMOP CDM. Mapping to a CDM harmonizes data query formats across various data sources. Specifically, data from EHR legacy systems, including Allscripts Touchworks (pre- 2021 for all clinical data) and GE Centricity (pre-2021 for billing) were combined with data from Epic (post-2021). These data were converted to the OMOP CDM on a nightly basis via an Apache Spark job. Structured data with available concept codes were directly converted from the source codes (International Disease Classification [ICD], Current Procedural Terminology [CPT], Healthcare Common Procedure Coding System [HCPCS], or Logical Observation Identifiers Names and Code [LOINC]) to the Athena concept vocabulary.^16^ Currently, the UTHealth OMOP CDM dataset includes data on the conditions, measurements, procedures, drug exposures, and clinical narratives of 6.5 million patients.

### Cohort construction

We constructed 3 cohorts of patients (pancreatic cancer and 2 types of STS) in both the IQVIA Oncology EMR and UTHealth OMOP CDM datasets. Because 60% of STS diagnoses occur in the extremities, 10% occur in the trunk, and 15% occur in the retroperitoneum, we created 2 STS cohorts, STS-TE and STS-AR, in each dataset.^14^ For each disease, a list of ICD-10 diagnosis codes was first determined by clinical experts (Supplementary Data 1). In real-world datasets, ICD codes may not capture patients with a true STS diagnosis. To address this challenge, we further refined our strategies to build cohorts of actual STS cases. In the IQVIA dataset, we used both ICD-10 codes and sarcoma histology results to retrieve STS patients. In the UTHealth OMOP CDM dataset, we first retrieved potential STS patients using the ICD-10 codes and then confirmed that the keyword “sarcoma” appeared in the patient’s clinical notes.

### Computation strategy

Our computation strategy included significant diagnosis pair mining and disease trajectory visualization. Figure 1 shows an overview of the computation strategy.

**Figure 1.**
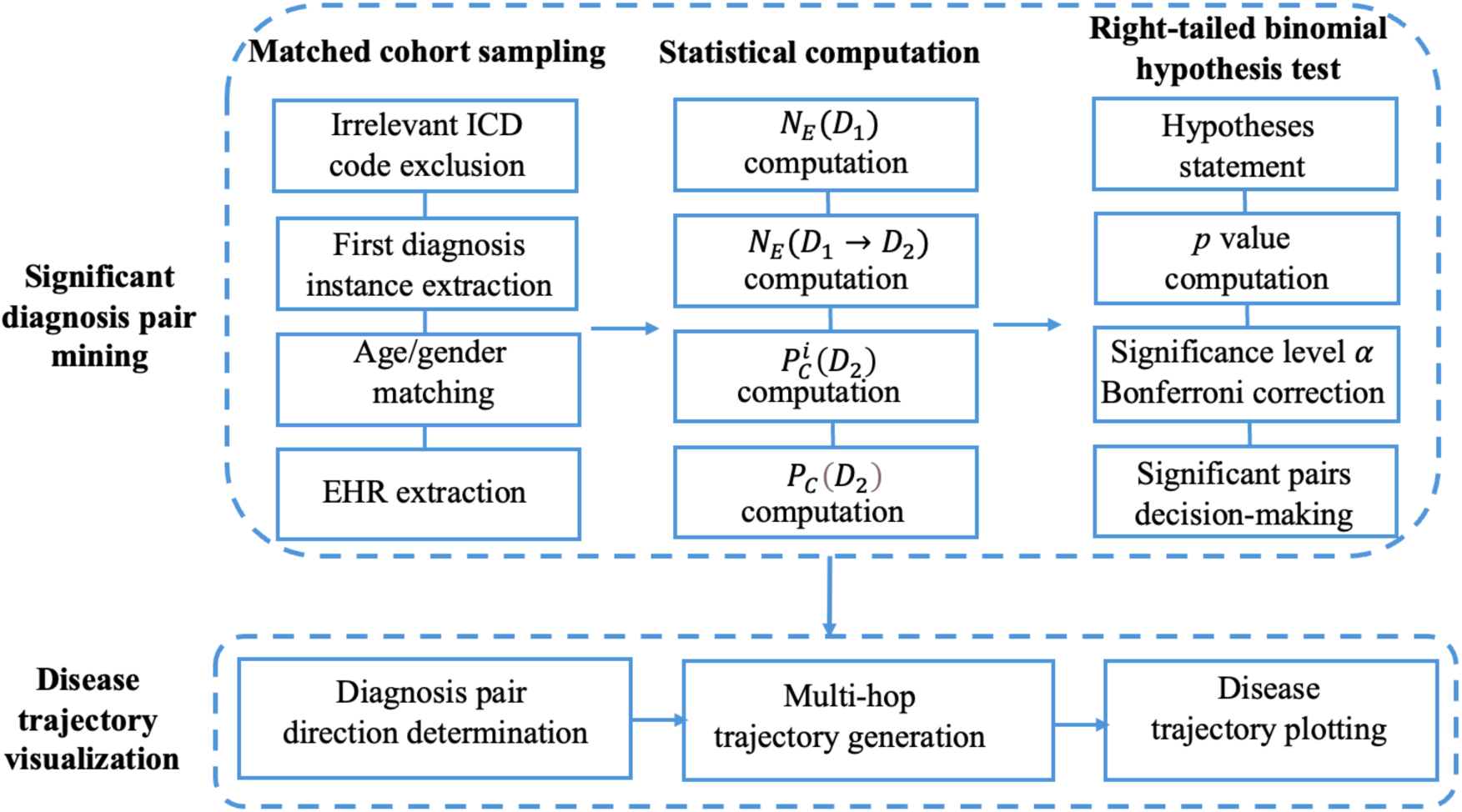
Overview of the computation strategy. EHR, electronic health record; ICD, International Classification of Diseases.

### Significant diagnosis pair mining

In identifying significant diagnosis pairs, our primary goal was to find the pair (*D*_1_, *D*_2_), in which the conditional probability *P_e_*(*D*_2_|*D*_1_) in patients with the cancer type (i.e., pancreatic cancer, STS-TE, or STS-AE) was higher than the marginal probability *P_c_*(*D*_2_) in the general population. This was achieved through matched cohort sampling, probability computation, and a right-tailed binomial hypothesis test.

#### Matched cohort sampling

To speed up computation, we extracted the first 4 digits of the full ICD-10 codes and excluded the irrelevant ICD-10 codes starting with “O__”, “P__”, “R__”, “S__”, “T__”, “V__”, “W__”, “X__”, “Y__”, and “Z__”. We retained only the first diagnosis of for any disease occurred in the cancer cohort and obtained all possible diagnosis pairs. Given a diagnosis pair (*D*_1_, *D*_2_), we defined the exposed group (*E*) and the comparison group (*C*). The exposed group contained patients diagnosed with both the cancer type (i.e., pancreatic cancer, STS-TE, or STS-AE) and *D*_1_. We extracted all diagnosis records starting from the diagnosis of *D*_1_ through the next 3 years in order to calculate *P_E_*(*D*_2_|*D*_1_). Each patient in the exposed group was matched to individuals from the entire IQVIA dataset according to age and gender to create a comparison group. We then extracted diagnosis records for these matched individuals within 3 years of their matching point. For example, we matched a female who was diagnosed with D1 at 60 in the exposed group to the females who were diagnosed with any disease at 60. We then extracted all diagnosis records of the matched patients within 3 years in the IQVIA. We pre-calculate the temporal diagnosis pair numbers across all 3 cancer types, and the average time interval for each pair. We found that there were 607,366 temporal diagnosis pairs in total and 87.3% of pairs occurred within a 3-year period. This justifies the use of the 3-year time window.

Notably, the patients in the exposed group had to be diagnosed with *D*_1_ but did not need to have a diagnosis of *D*_2_, as the aim of collecting data from the exposed group was to calculate the probability *P*_*E*_(*D*_2_|*D*_1_) to compare with the marginal probability *P*_*C*_(*D*_2_). Patients in the comparison group did not have to have a diagnosis of *D*_1_ or *D*_2_.

#### Statistical computation

Given the diagnosis pair (*D*_1_, *D*_2_) and the corresponding exposed group (*E*) and comparison group (*C*), *N*_*E*_(*D*_1_) represents the number of patients who were diagnosed with *D*_1_ in the exposed group, and *N*_*E*_(*D*_1_ → *D*_2_) represents the number of patients who were diagnosed with *D*_1_ and diagnosed with *D*_2_ within 3 years. *P*_*C*_(*D*_2_) represents the probability of a patient in the comparison group being diagnosed with *D*_2_ (Figure 2).

**Figure 2.**
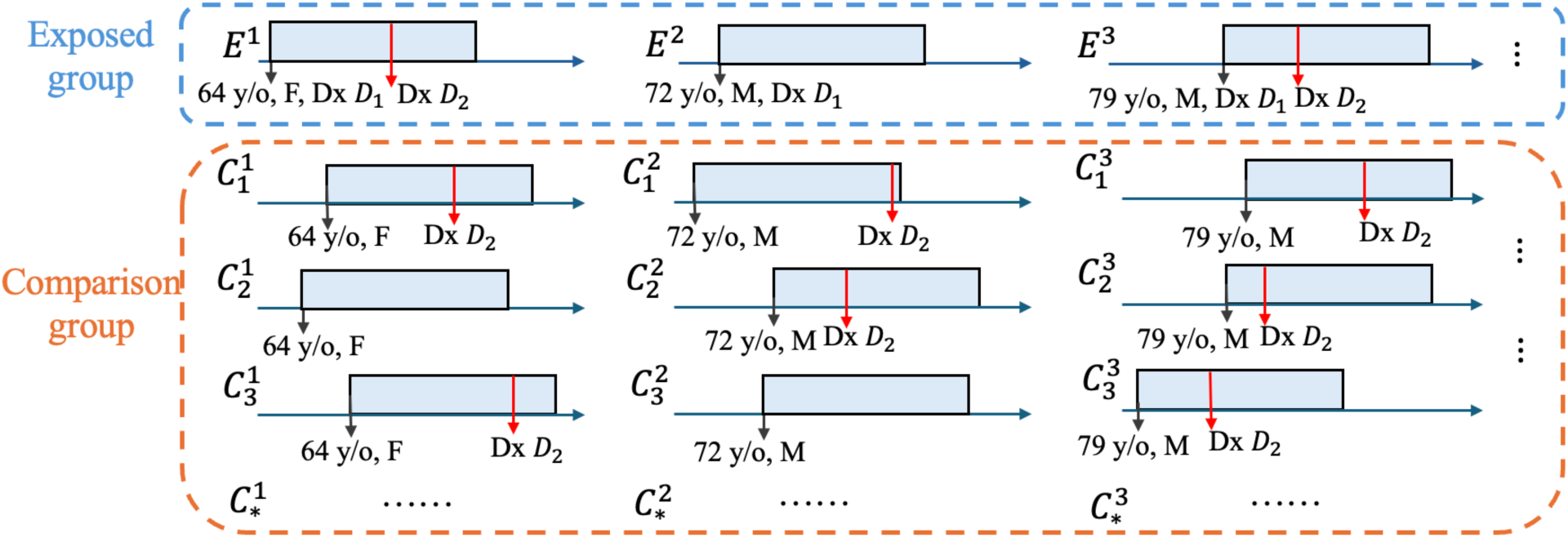
Matched cohort sampling and probability computation. The blue dashed box represents the exposed group, consisting of 3 example patients (*E*^1^, *E*^2^, *E*^3^). All of these patients were diagnosed with *D*_1_ and may or may not have been diagnosed with *D*_2_. The orange box represents the comparison group, containing multiple patients who were age- and gender-matched to the patients in the exposed group (*C*_*_^1^, *C*_*_^2^, *C*_*_^3^). It is important to note that although only 3 matched examples displayed, the comparison group includes all patients meeting the matching criteria from the entire IQVIA dataset. The superscript ∗ denotes the index of the patient in the exposed group. F, female; M, male; y/o, years old; Dx, diagnosis

For each patient in the exposed group, *E*^*i*^, *P*^*i*^ (*D*_2_) represents the probability of being diagnosed with *D*_2_ across all patients in that exposed patient’s comparison group. The overall probability *P_c_*(*D*_2_) is computed as the average of *P*^*i*^ (*D*_2_) across all patients in the exposed group. If fewer than 2,000 patients were matched for any given *E*^*i*^, the match was deemed unrepresentative and the *E*^*i*^ was excluded from the exposed group when statistically analyzing the (*D*_1_, *D*_2_) pair.

To optimize the computation process, we performed a pre-filtering step, keeping only the diagnosis pairs that were present in more than 0.75% of patients with the cancer of interest. This percentage could be adjusted based on computational capacity

#### Right-tailed binomial hypothesis test

To determine whether the pair (*D*_1_, *D*_2_) was significant, we conducted a right-tailed binomial hypothesis test. The hypotheses were:

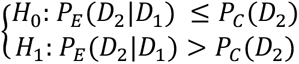

We used a binomial distribution to model *N*_*E*_(*D*_1_ → *D*_2_). In this case, the number of trials is *N*_*E*_(*D*_1_), the success probability under the null hypothesis is *P*_*C*_(*D*_2_), and the observed number of successes is *N*_*E*_(*D*_1_ → *D*_2_). The *p* value is the observed probability when *N*_*E*_(*D*_1_ → *D*_2_) is greater than *P*_*C*_(*D*_2_) ∗ *N*_*E*_(*D*_1_). We first set the significance level *α* to 0.05. Because there were thousands of diagnosis pairs, we performed the Bonferroni correction^17^ to adjust the significance level to control the overall type I error rate across multiple comparisons. The adjusted significance level *α*′ was computed, with *α*′ = *α*/*m*, where *m* is the number of diagnosis pairs. Then, we computed *p* = *P*(*N*_*E*_(*D*_1_ → *D*_2_) > *P*_*C*_(*D*_2_) ∗ *N*_*E*_(*D*_1_)). If *p* < *α*′, *H*_0_ was rejected, i.e., if *N*_*E*_(*D*_1_ → *D*_2_) was significantly greater than *P*_*C*_(*D*_2_) ∗ *N*_*E*_(*D*_1_) and thus *P*_*E*_(*D*_2_|*D*_1_) > *P*_*C*_(*D*_2_), the pair (*D*_1_, *D*_2_) was significant.

### Disease trajectory visualization

Disease trajectory visualization was achieved through the determination of the directionality of the diagnosis pairs, multi-hop trajectory generation, and disease trajectory plotting. A “hop” refers to a progression from one disease to another (i.e., a new diagnosis).

#### Determination of the directionality of diagnosis pairs

The diagnosis pairs (*D*_1_, *D*_2_) and (*D*_2_, *D*_1_) both could have been identified as significant. In this case, we needed to determine the overall directionality of that diagnosis pair. *N*_*E*_(*D*_1_ → *D*_2_) denotes the number of patients diagnosed with *D*_1_ and then diagnosed with *D*_2_, and *N*_*E*_(*D*_2_ → *D*_1_) denotes the number of patients diagnosed with *D*_2_ and then diagnosed with *D*_1_. We set *N*_*E*_(*D*_1_, *D*_2_) = *N*_*E*_(*D*_1_ → *D*_2_) + *N*_*E*_(*D*_2_ → *D*_1_). Then, we compared *N*_*E*_(*D*_1_ → *D*_2_) and *N*_*E*_(*D*_2_ → *D*_1_) with 0.5 ∗ *N*_*E*_(*D*_1_, *D*_2_), and performed the right-tailed binomial hypothesis test to determine the overall directionality of the disease trajectory. If the directionality from *D*_1_ → *D*_2_ was significant, it indicated that the diagnosis of *D*_1_ preceded that of *D*_2_, establishing a directional link from *D*_1_ to *D*_2_ in the disease trajectory. If the directionality from *D*_2_ → *D*_1_ was significant, it indicated that the diagnosis of *D*_2_ preceded that of *D*_1_, establishing a directional link from *D*_2_ to *D*_1_ in the disease trajectory. If neither directionality was significant, it suggested that *D*_1_ and *D*_2_ were co-occurring.

#### Multi-hop trajectory generation

Long trajectories with multiple hops were constructed by sequentially concatenating 1-hop (diagnosis pair) trajectories with directionalities. For instance, if both *D*_1_ → *D*_2_ and *D*_2_ → *D*_3_ were significant, *D*_1_ → *D*_2_ → *D*_3_ formed a 2-hop trajectory with 3 nodes. If this trajectory was included in the EHR data of patients with the disease of interest, we classified *D*_1_ → *D*_2_ → *D*_3_ as a valid 2- hop trajectory. Once all 2-hop trajectories were identified, we concatenated them with the 1-hop trajectories to generate potential 3-hop trajectories. These 3-hop trajectories were then filtered so that only those present in at least 1 patient’s EHR data were retained. We continued this process iteratively, expanding the trajectory length until no additional trajectories were identified.

#### Disease trajectory plotting

After obtaining the diagnosis pairs with and without directionality, we used Gephi 0.10,^18^ an open-source software platform designed for visualizing and analyzing large networks and graphs, to visualize the disease trajectories.

### Trajectory validation

We systematically validated trajectories with 1 hop, 2 hops, and 3 hops with and without directionality using patient data encompassing all 3 types of cancer from the UTHealth OMOP CDM. Specifically, we kept only the first diagnosis date of any disease occurred in all patients with a pancreatic cancer, STS-TE, or STS-AR diagnosis. We then compared the temporally ordered diagnoses of cancer patients with the discovered trajectories, and the overlapping trajectories were regarded as validated. For example, in the 2-hop trajectory of unspecified anemia (D64.9), unspecified iron-deficiency anemia (D50.9), and STS-TE, if a patient had temporally ordered diagnoses of these diseases, then the trajectory was validated.

## RESULTS

### Patient characteristics

Table 1 shows the demographic characteristics of the cancer cohorts in the IQVIA and UTHealth OMOP CDM. The IQVIA cohort had approximately 5 times more patients than the UTHealth OMOP CDM cohort.

**Table 1.**
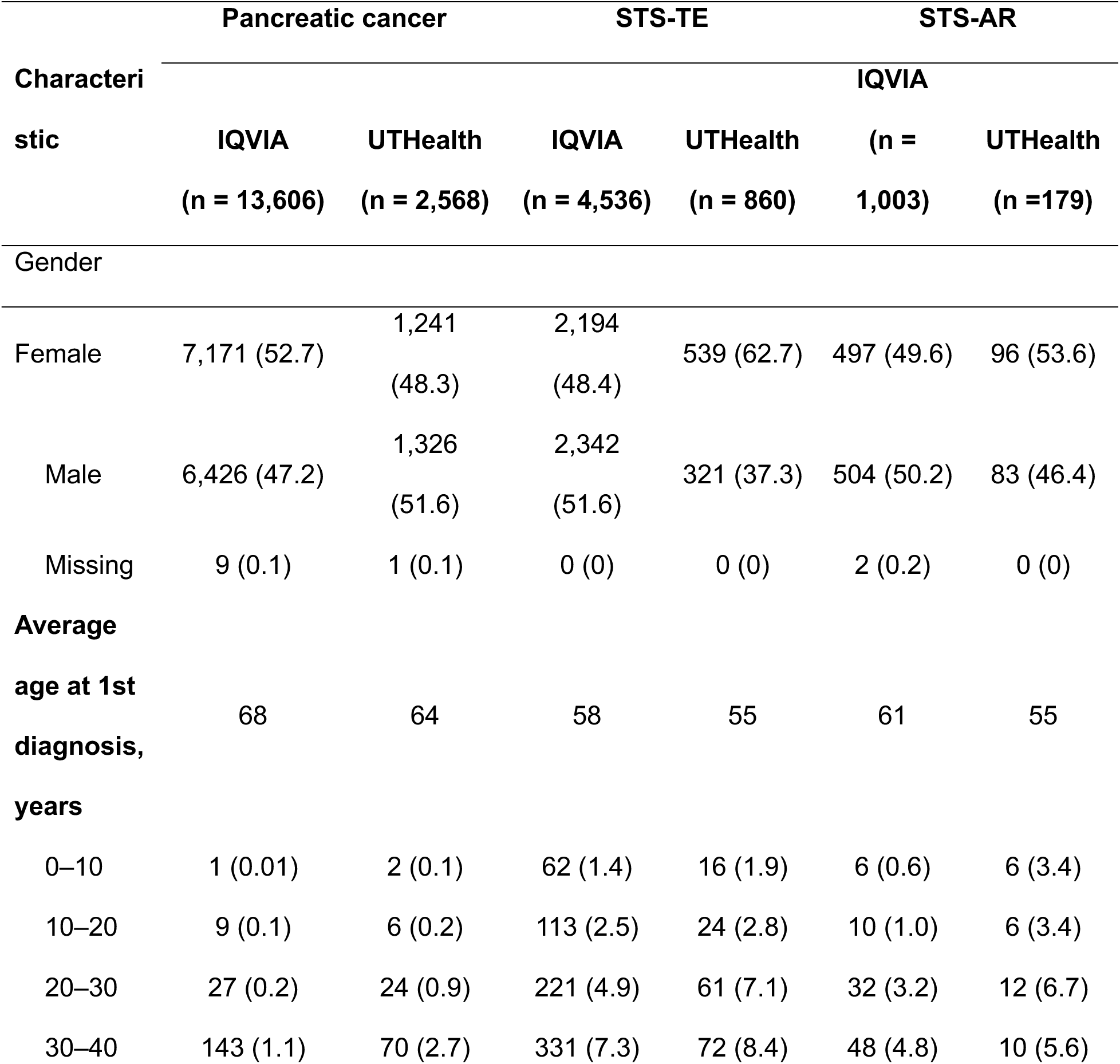
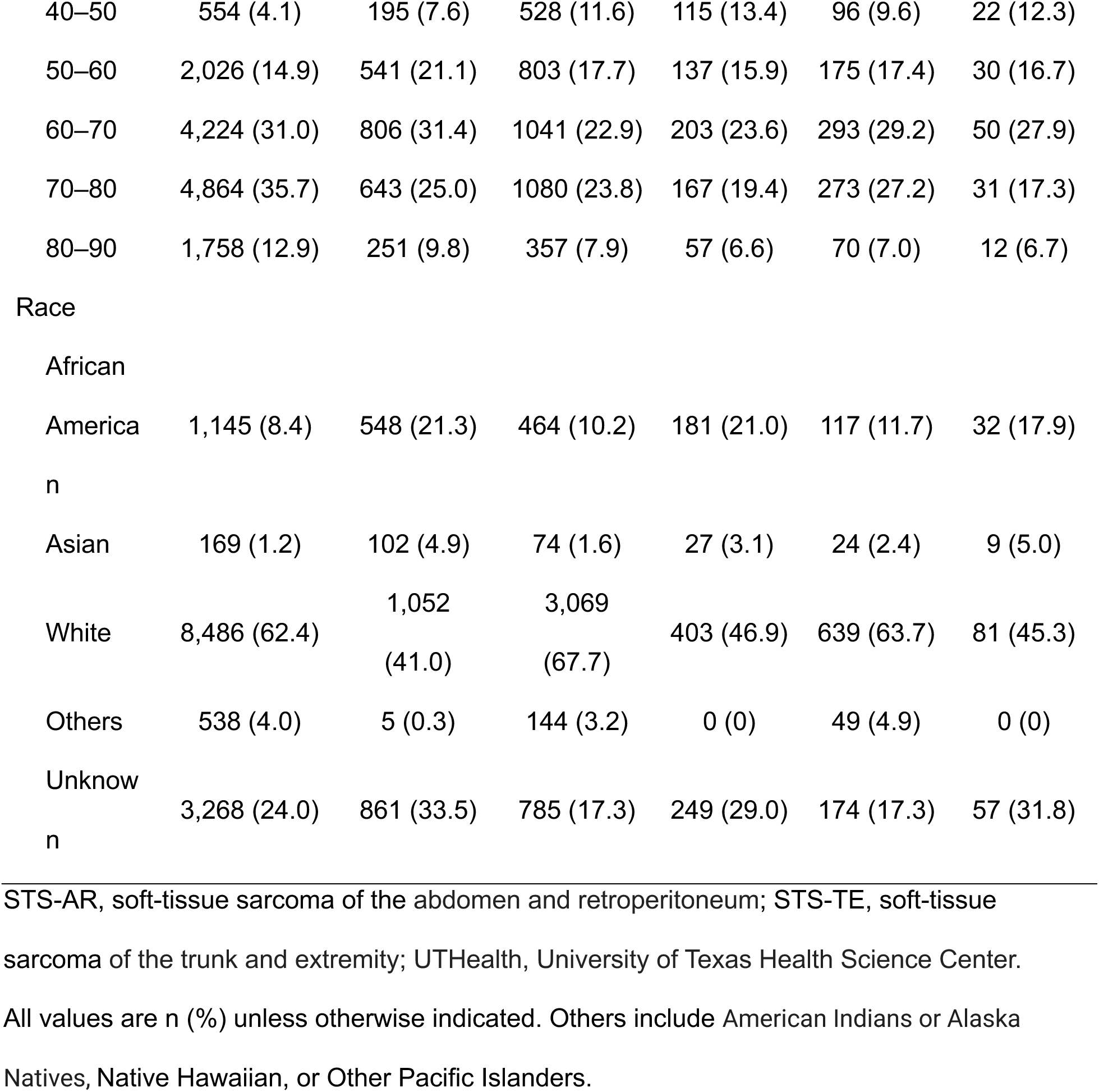
Demographic characteristics of patients by cancer type and dataset (IQVIA vs. UTHealth OMOP CDM).

### Significant diagnosis pairs

Table 2 shows the relevant values for calculating significant diagnosis pairs for the 3 cancer categories. Among the 13,606 pancreatic cancer patients, 389,919 ICD-10 diagnosis pairs were initially found. After pre-filtering, only 469 diagnosis pairs remained. The Bonferroni-corrected significance level was set to 1.28 × 10^−7^ with *α*′ = *α*/*N*. We then conducted a right-tailed binomial hypothesis test, identifying 266 diagnosis pairs as significant. Regarding directionality, 16 diagnosis pairs were found to have no directionality (i.e., were co-occurring), and 250 diagnosis pairs were found to be directional. Among those 250 pairs, 39 pairs followed pancreatic cancer, 46 pairs preceded pancreatic cancer, and 165 pairs contained no pancreatic cancer diagnoses. There were 130 significant pairs, with 124 being directional for STS-TE, and 118 significant pairs, with 113 being directional for STS-AR. Tables S1, S6 and S11 of the Supplementary Data 2 contain the significant pairs for each type of cancer.

**Table 2.**
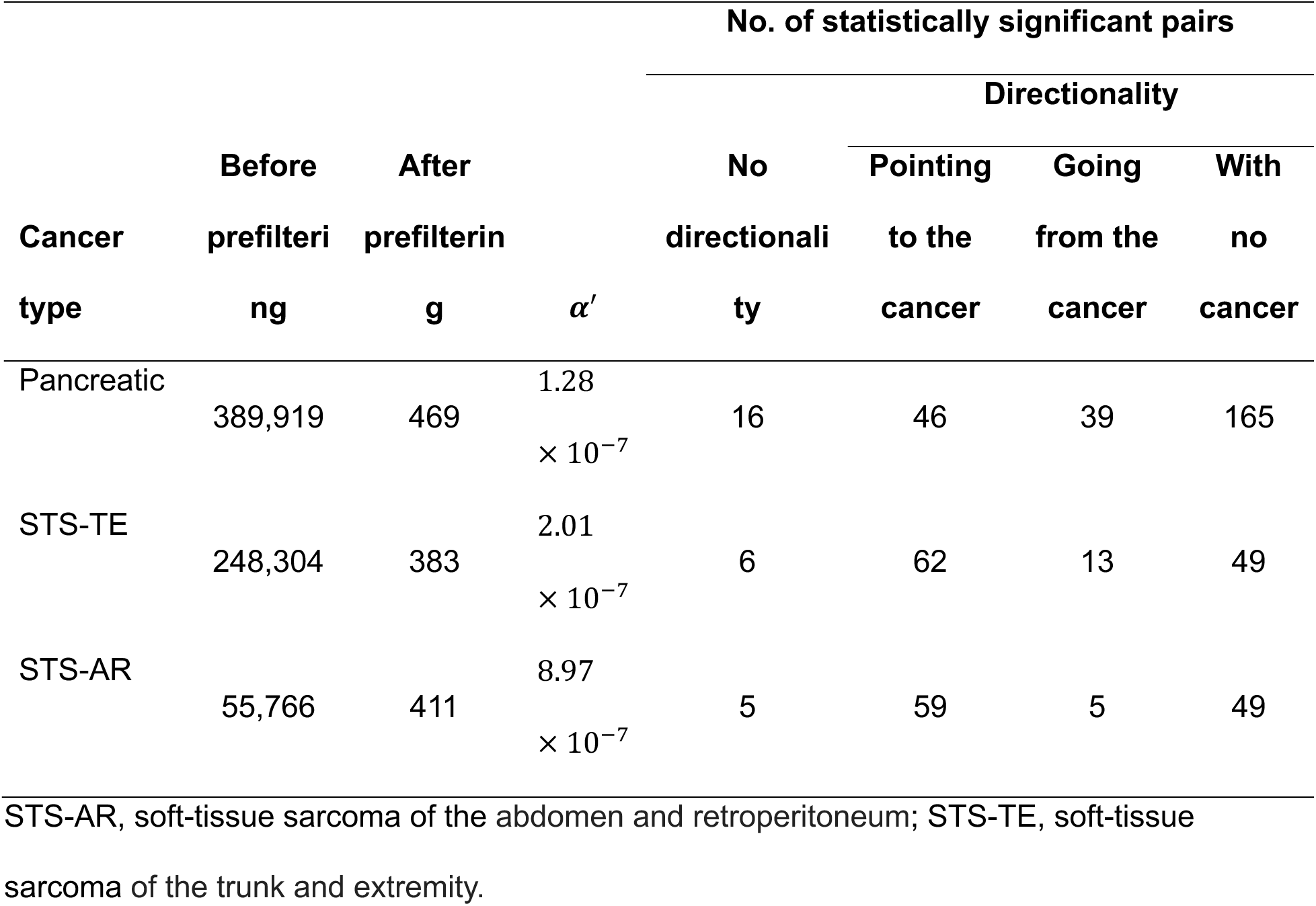
Calculation of diagnosis pairs.

### Disease trajectories

Figure 3, Figure 4, and Figure 5 show the significant disease trajectories of pancreatic cancer, STS-TE, and STS-AR, respectively. In terms of diagnosis pairs with the highest edge strength (i.e., most common trajectories), the top 3 diagnoses that preceded pancreatic cancer were primary hypertension (I10) with 954 patients accounting for 7% of all pancreatic patients in IQVIA, hyperlipidemia (E78.5) (595, 4.4%), and type 2 diabetes mellitus (E11.9) (553, 4.1%). The top 3 diagnoses that followed pancreatic cancer were dehydration (E86.0) (1828, 13.4%), neoplasm-related pain (G89.3) (1301, 9.6%), and Secondary malignant neoplasm of liver and intrahepatic bile ducts (C78.7) (1197, 8.8%). A total of 43 disease diagnoses preceded both STS-TE and STS-AR, accounting for 69.4% and 72.9% of prior diagnoses, respectively. The top 3 diagnoses that preceded STS-TE were primary hypertension (I10) (285, 6.3%); pain in a joint (M25.5) (219, 4.8%); and pain in a limb, hand, foot, finger, or toe (M79.6) (175, 3.9%). The top 3 diagnoses that preceded STS-AR were primary hypertension (I10) (69, 6.9%), unspecified anemia (D64.9) (42, 4.2%), and pain in a joint (M25.5) (41, 4.1%). A total of 2 disease diagnoses followed both STS-TE and STS-AR, accounting for 15.4% and 40.0% of prior diagnoses, respectively. The top 3 diagnoses that followed STS-TE were secondary malignant neoplasm of the lung (C78.0) (238, 5.2%), neoplasm-related pain (G89.3) (229, 5%), and agranulocytosis secondary to cancer chemotherapy (D70.1) (162, 3.6%). The top 3 diagnoses that followed STS-AR were malignant neoplasm of connective and soft tissue (C49.9) (101, 10%), unspecified neoplasm-related pain (G89.3) (58, 5.8%), and secondary malignant neoplasm of the lung (C78.0) (38, 3.8%).

**Figure 3.**
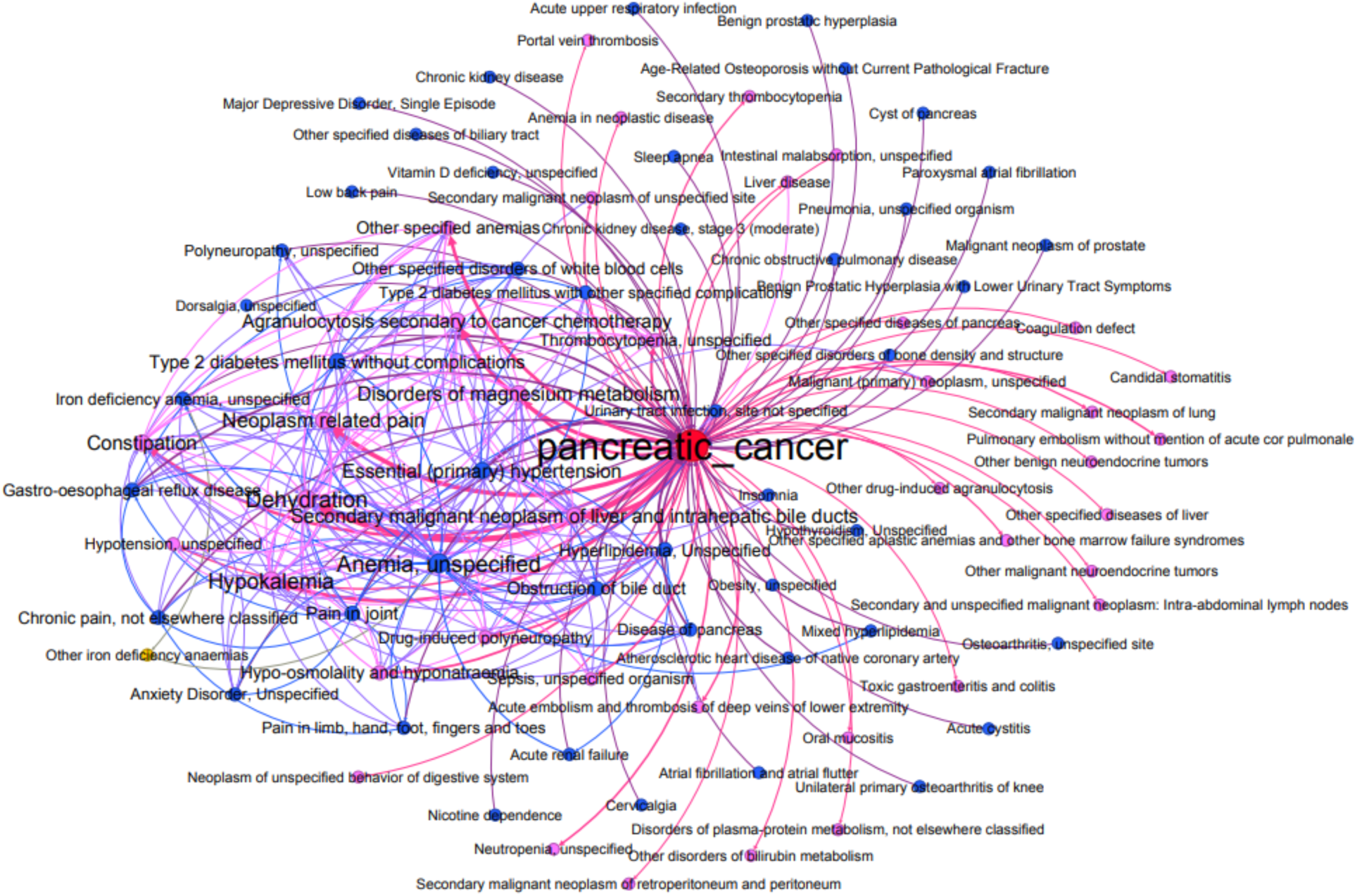
Disease trajectories of patients with pancreatic cancer. The thickness of each trajectory (i.e., line) represents the number of patients associated with that trajectory; thicker arrows represent trajectories containing more patients. The nodes in the trajectories are categorized as follows: red represents pancreatic cancer; blue represents nodes that point to pancreatic cancer, indicating that the diagnosis nodes precede the pancreatic cancer diagnosis; pink refers to nodes pointed away from pancreatic cancer, indicating that the diagnosis occurred after the pancreatic cancer diagnosis; and yellow indicates that a node was not linked to pancreatic cancer.

**Figure 4.**
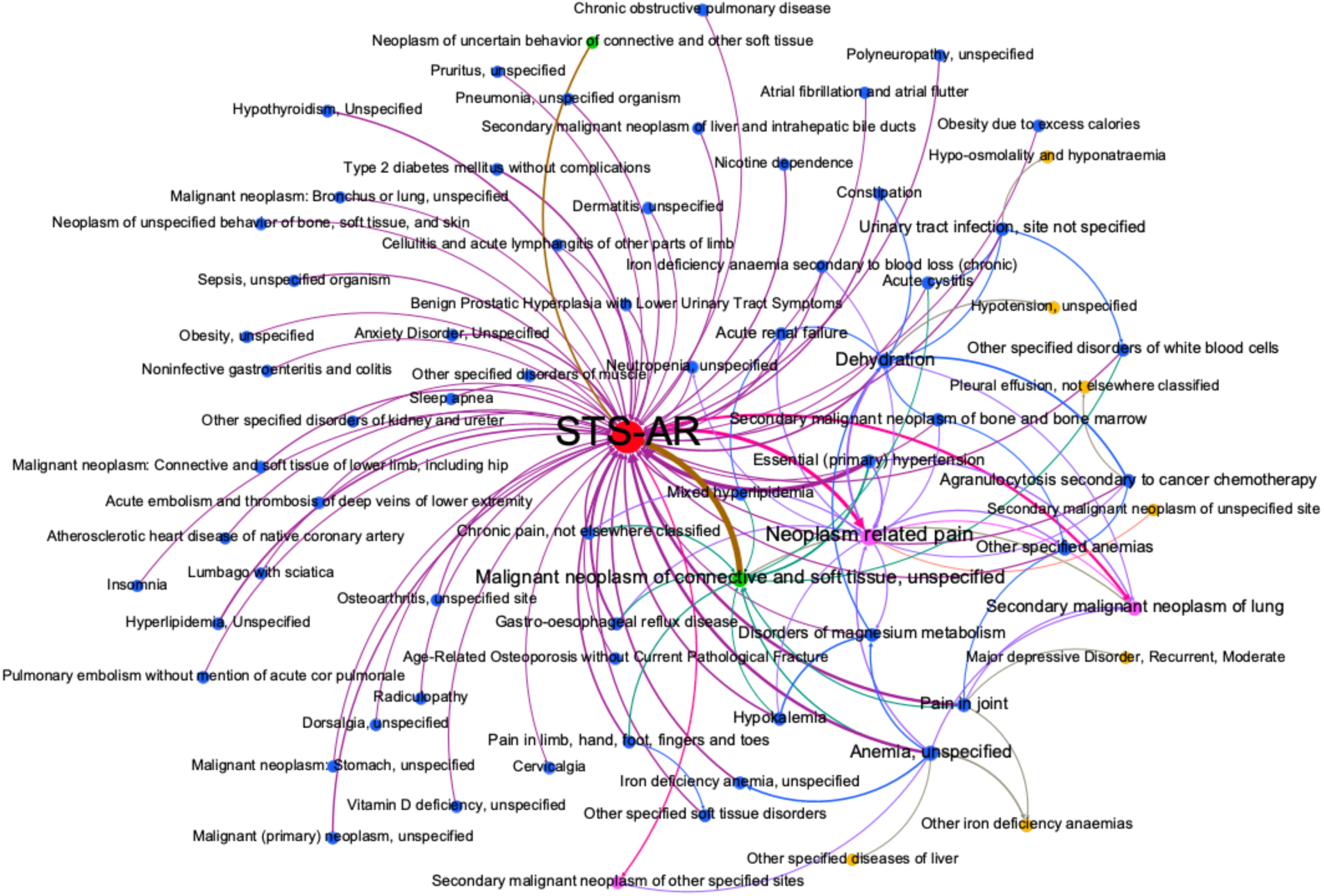
Disease trajectories of soft-tissue sarcoma of the abdomen and retroperitoneum (STS-AR). The thickness of each trajectory (i.e., line) represents the number of patients associated with that trajectory; thicker arrows represent trajectories containing more patients. The disease nodes in the trajectories are categorized as follows: red represents STS- AR; blue represents nodes that point to STS-AR, indicating that the diagnosis nodes precede the STS-AR diagnosis; pink refers to nodes pointed away from STS-AR, indicating that the diagnosis occurred after the STS-AR diagnosis; green represents nodes linked to STS-AR with no directionality, indicating that the disease co-occurred with STS-AR; and yellow indicates that a node was not linked to STS-AR.

**Figure 5.**
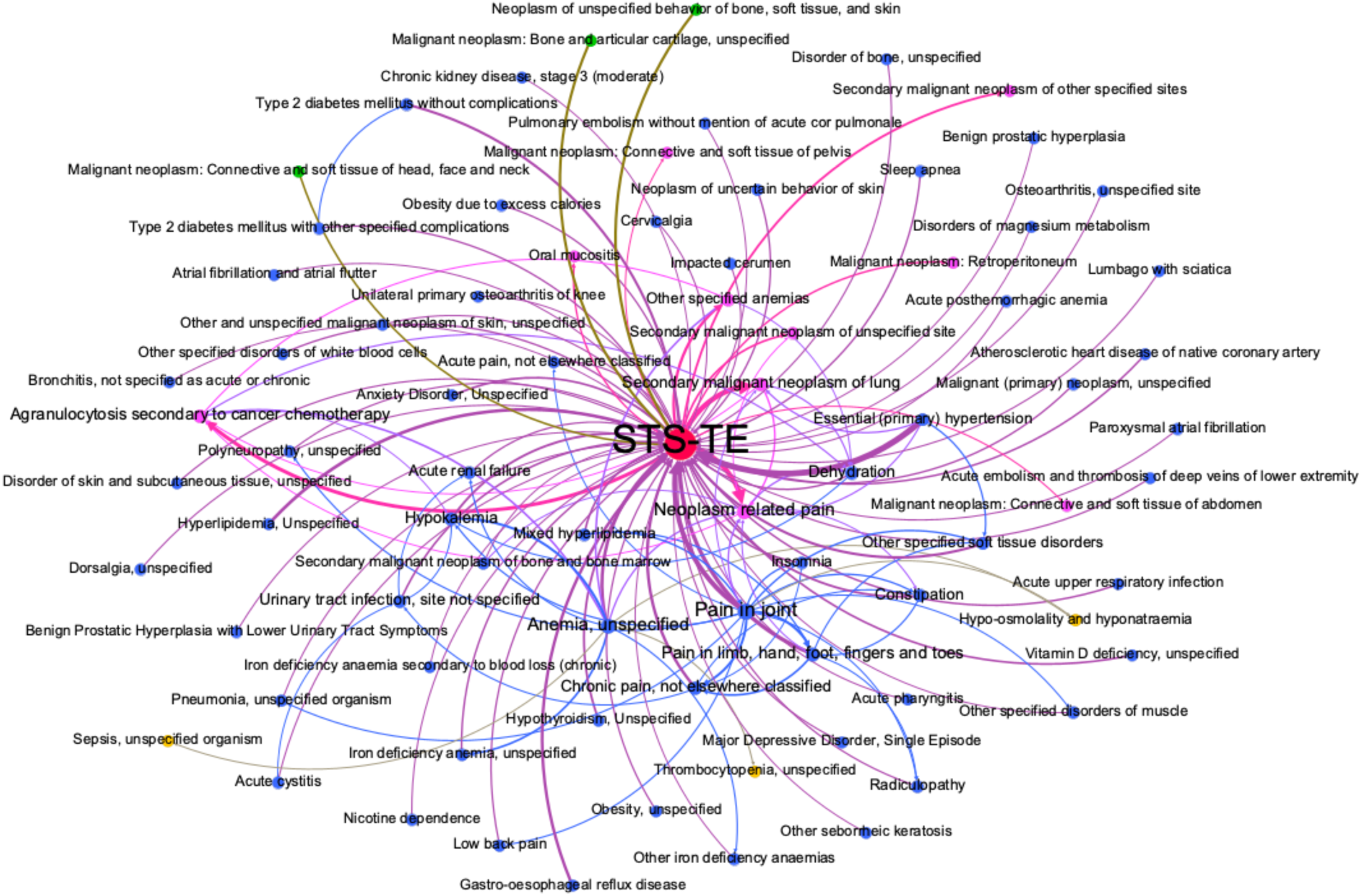
Disease trajectories of soft-tissue sarcoma of the trunk and extremity (STS-TE). The thickness of each trajectory (i.e., line) represents the number of patients associated with that trajectory; thicker arrows represent trajectories containing more patients. The disease nodes in the trajectories are categorized as follows: red represents STS-TE; blue represents nodes that point to STS-TE, indicating that the diagnosis nodes precede the STS-TE diagnosis; pink refers to nodes pointed away from STS-TE, indicating that the diagnosis occurred after the STS-TE diagnosis; green represents nodes linked to STS-AR with no directionality, indicating that the disease co-occurred with STS-TE; and yellow indicates that a node was not linked to STS-TE.

Table 3 shows the number of long trajectories for each cancer type. Figure 6 shows the example disease trajectories that preceded a pancreatic cancer, STS-TE, or STS-AR diagnosis found in most patients. For example, type 2 diabetes, gastroesophageal reflux disease and anemia occurred before pancreatic cancer. Hyperlipidemia occurred before STS-TE. Figure 7 shows the example disease trajectories following pancreatic cancer, STS-TE, or STS-AR diagnosis found in most patients. Supplementary Data 2 (Tables S2-S14 excluding S6 and S11) contains the detailed data of the long trajectories (i.e., those with multiple hops) discovered from the IQVIA Oncology EMR for pancreatic cancer, STS-AR, and STS-TE. For example, hypertension occurred before STS.

**Figure 6.**
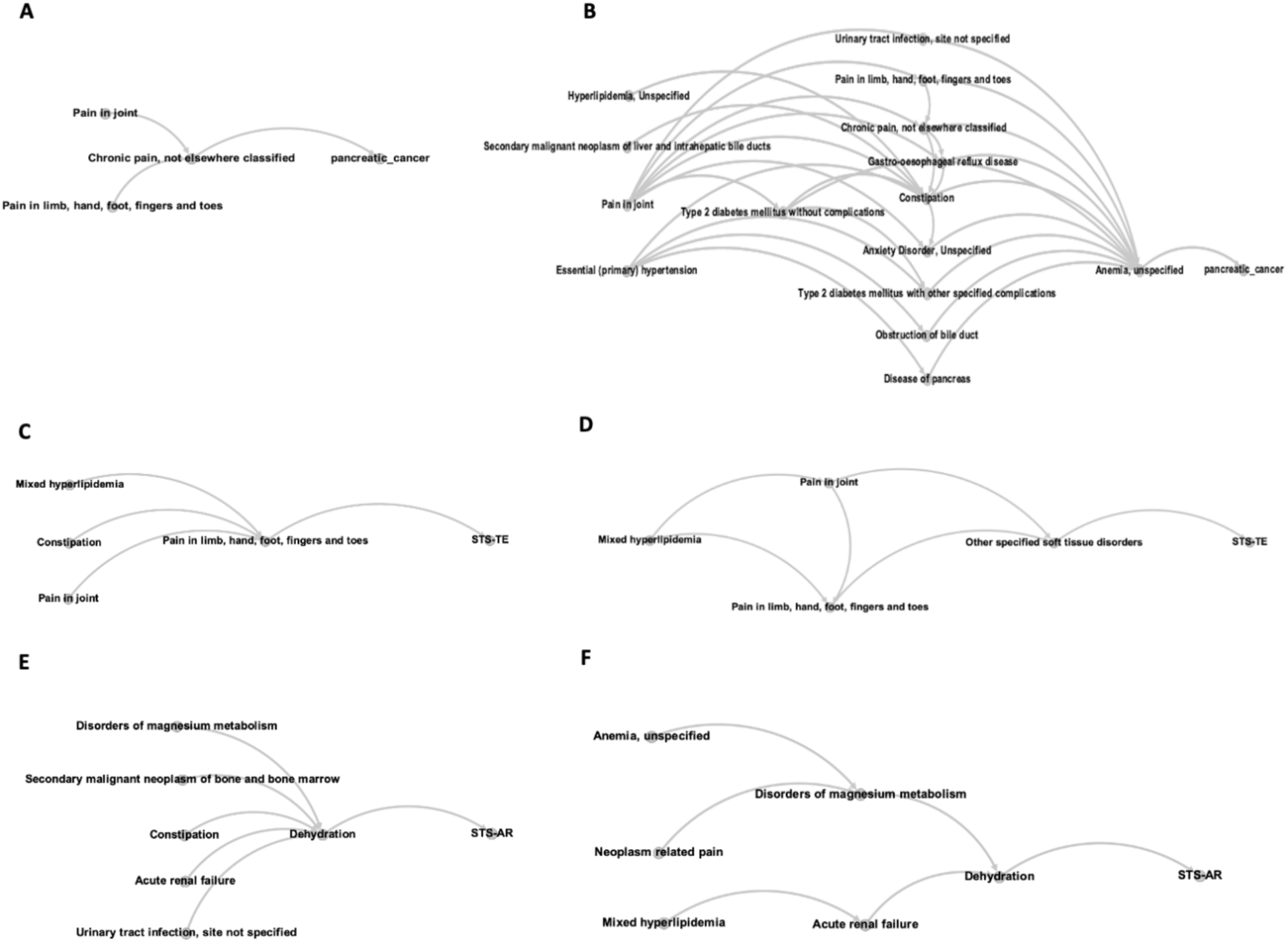
Possible disease trajectories before pancreatic cancer, STS-TE and STS-AR diagnoses. A) Trajectories with 2 hops before a pancreatic cancer diagnosis. B) Trajectories with 3 hops before a pancreatic cancer diagnosis. C) Trajectories with 2 hops before an STS-TE diagnosis. D) Trajectories with 3 hops before an STS-TE diagnosis. E) Trajectories with 2 hops before an STS-AR diagnosis. F) Trajectories with 3 hops before an STS-AR diagnosis.

**Figure 7.**
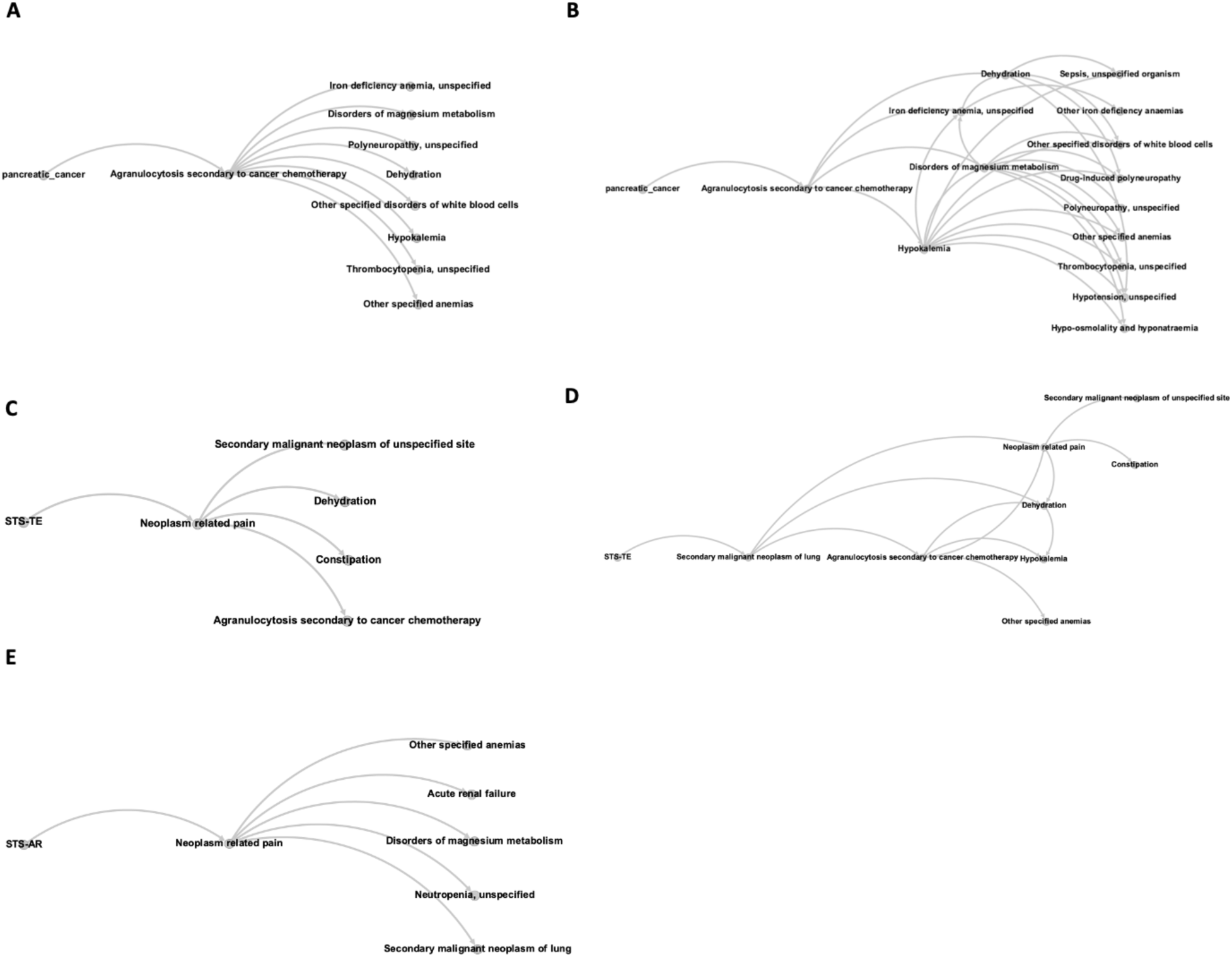
Possible disease trajectories after pancreatic cancer, STS-TE, and STS-AR diagnoses. A) Trajectories with 2 hops after a pancreatic cancer diagnosis. B) Trajectories with 3 hops after a pancreatic cancer diagnosis. C) Trajectories with 2 hops after an STS-TE diagnosis. D) Trajectories with 3 hops after an STS-TE diagnosis. E) Trajectories with 2 hops after an STS-AR diagnosis. No trajectories with 3 hops after an STS-AR diagnosis were identified.

**Table 3.**
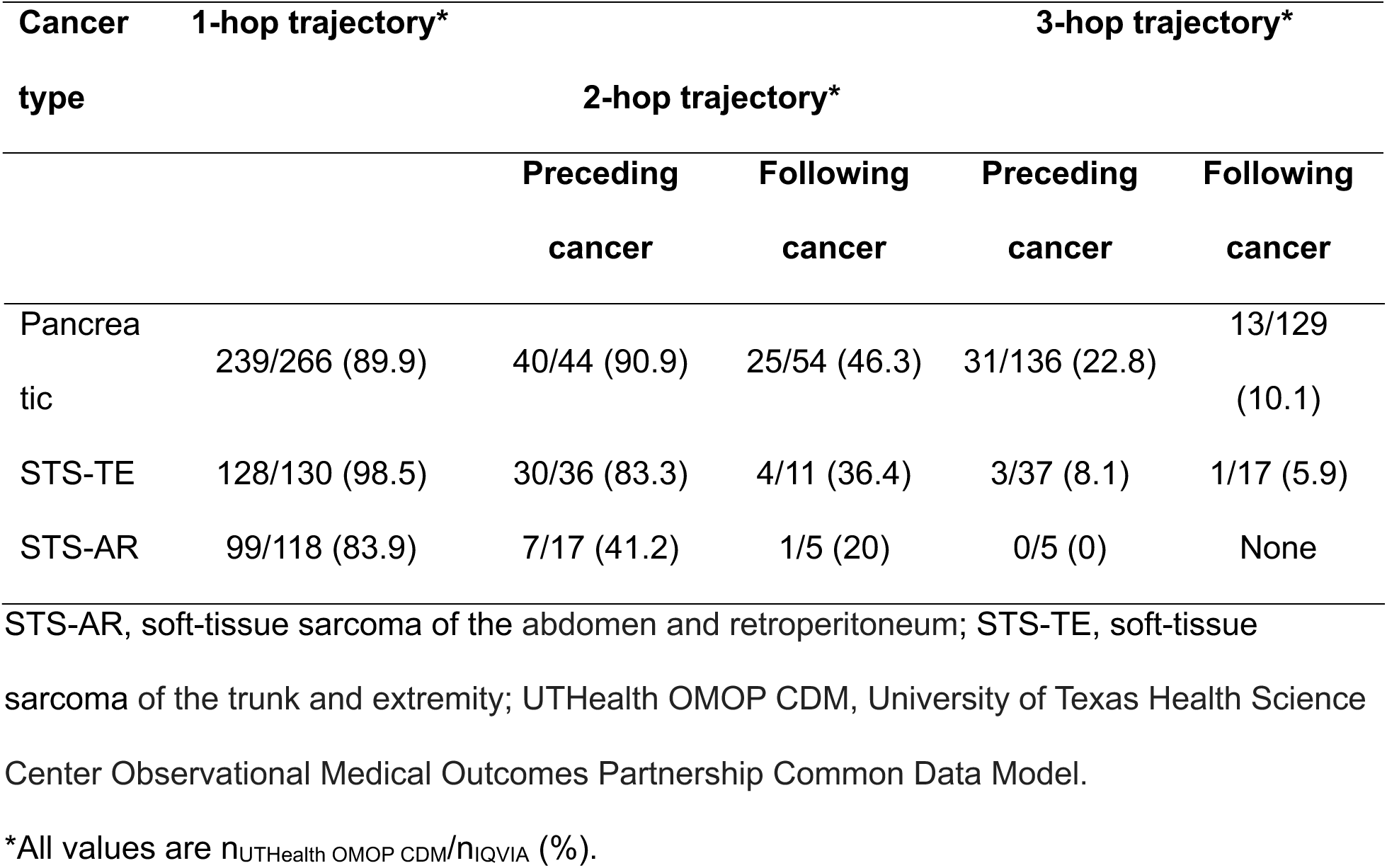
Validation results of pairs with directionalities and long trajectories.

### Trajectory validation

We systematically validated the significant disease trajectories with local data from the UTHealth OMOP CDM dataset to demonstrate the accuracy of our approach. Table 3 presents the validation results from the UTHealth OMOP CDM dataset for diagnosis pairs and long (i.e., 2- and 3-hop) trajectories identified in the IQVIA dataset. The results showed that 89.9%, 98.5%, and 83.9% of 1-hop trajectories found in the IQVIA cohorts were also found in the pancreatic cancer, STS-TE, and STS-AR patient cohorts of the UTHealth OMOP CDM, respectively. More long disease trajectories were found preceding the cancer diagnoses than following the cancer diagnoses in the UTHealth OMOP CDM cohorts. Specifically, trajectories with 2 hops preceding a pancreatic cancer, STS-TE, and STS-AR diagnosis were identified in 90.9% (vs. 46.3%), 83.3% (vs. 36.4%), and 41.2% (vs. 20%), respectively, of the UTHealth OMOP CDM cohorts. Longer disease trajectories (i.e., those with 3 hops) preceding and following cancer diagnoses were less covered. Trajectories with 3 hops preceding a pancreatic cancer or STS-TE diagnosis were covered by 22.8% and 8.1%, and trajectories with 3 hops following a pancreatic cancer or STS-TE diagnosis were covered by 10.1% and 5.9%, respectively, of patients in the UTHealth OMOP CDM dataset cohorts.

## DISCUSSION

In this study, we extracted signature disease trajectories from the IQVIA Oncology EMR for 3 rare cancers, which we subsequently validated using the UTHealth OMOP CDM dataset. Our identification of signature disease trajectories demonstrated the value of rendering real-world big data to better understand rare cancer disease patterns; 13,606 pancreatic cancer, 4,536 STS-TE, and 1,003 STS-AR patients in the IQVIA Oncology EMR and 2,568, 860, and 179 patients, respectively, in the UTHealth OMOP CDM were included in our study. This aggregation of rare cancer cases, along with the remaining 1.2 million cancer patients in the IQVIA oncology dataset that were included in the comparison groups, provided a rich and robust foundation for identifying signature disease trajectories. Our findings included 266 significant diagnosis pairs for pancreatic cancer, 130 significant pairs for STS-TE, and 118 significant pairs for STS-AR. In addition, we discovered 44 2-hop (i.e., 2-progression) and 136 3-hop trajectories before a pancreatic cancer diagnosis, 36 2-hop and 37 3-hop trajectories before an STS-TE diagnosis, and 17 2-hop and 5 3-hop trajectories before an STS-AR diagnosis. Meanwhile, we found 54 2- hop and 129 3-hop trajectories following a pancreatic cancer diagnosis, 11 2-hop and 17 3-hop trajectories following an STS-TE diagnosis, and 5 2-hop and 0 3-hop trajectories following an STS-AR diagnosis.

Our study offers 3 key contributions to understanding disease progression patterns. First, our approach is tailored to extract signature trajectories for a specific disease of interest. In our study, we identified distinct disease trajectories for 3 rare cancers. Notably, the two STS types exhibited some unique progression patterns. For example, essential (primary) hypertension (I10) progressed to other specified soft tissue disorders (M79.8) before a diagnosis of STS-TE (Table S7 of Supplementary Data 2, while this pattern was not significant in STS-AR (Table S12 of Supplementary Data 2). Mixed hyperlipidemia (E78.2) progressed to acute renal failure (N17.9), and dehydration (E86.0) before a diagnosis of STS-AR (Table S14 of Supplementary Data 2), while usually pain followed mixed hyperlipidemia before a diagnosis of STS-TE (Table S9 of Supplementary Data 2). Although we used the IQVIA Oncology EMR as the main data source, such an approach is generalizable to other diseases and real-world longitudinal patient records. Second, we systematically validated the discovered trajectories with local data, supporting the validity of the discovered trajectories. To our knowledge, this has not been done in previous studies. Third, the disease trajectories of pancreatic cancer, STS-TE, and STS-AR discovered in this study are potential resources for providers to deepen their understanding of the temporal progression of conditions preceding and following these rare cancers, further informing patient-care decisions.

Understanding disease trajectories is particularly critical in the rare cancer population, as identifying high-risk patients who may benefit from early surveillance or screening is challenging. Presently, high-risk surveillance for pancreatic cancer is typically reserved for patients with cystic lesions in their pancreas or those who have high familial and/or genetic risk.^16^ Similarly, STS screening is reserved for patients with high-risk genetic alterations such as *TP53*, *RB1*, and *NF1* mutations. Our findings suggest that key clinical features and diagnoses such as hyperlipidemia, pain in the joints, urinary tract infection, disorders of magnesium metabolism, acute renal failure, and dehydration may precede the diagnoses of pancreatic cancer and STS that can potentially serve as early markers of disease.

Furthermore, some of the significant diagnosis pairs identified in our study have been reported in other studies. However, longer trajectories were not reviewed in the existing literature. For example, new-onset diabetes has been associated with a higher risk of pancreatic cancer.^19^ A modest causal association between type 2 diabetes and pancreatic cancer was also established in a meta-analysis of 36 studies.^20^ Another study using the UK Biobank data established a possible relationship between gastroesophageal reflux disease and pancreatic cancer.^21^ Anemia, another diagnosis closely associated with pancreatic cancer in our study, has been found in 93% of patients presenting with pancreatic cancer.^22^ Sarcoma could be misdiagnosed as chronic hypertension ^23^ and in some sarcoma patients, hyperlipidemia was found in their medical history.^24, 25^

The early signs and symptoms of STS are often challenging to differentiate from those of benign conditions. Patients with palpable masses associated with pain or growth are often recommended to undergo further evaluation.^26^ However, patients with STS often experience significant delays in referral and treatment; one recent study suggested that patients can experience up to 14-month delays before a referral to a high-volume center.^27^ In this same study, pain alone was not enough for a patient to meet the criteria for referral to a sarcoma center, despite earlier research listing it as one of 5 criteria for urgent screening.^28^ Our data support these findings in earlier research, suggesting that pain in the joint or extremity may be an early marker of an STS-TE diagnosis. Similarly, STS-AR has vague signs and symptoms. Our 1- and 2-hop trajectories suggest that in patients with increasing abdominal girth or distention in conjunction with anemia and acute renal failure, clinicians should have a high index of suspicion for STS-AR.

Our work has the following limitations. First, the IQVIA dataset only spanned 8 years (2015 to 2023). If a patient was diagnosed with one of the diseases of interest after 2022, enough post-diagnosis data may not have been available from the 3-year period we used to calculate diagnosis pairs. Applying our method to longitudinal patient records over a longer duration is warranted to discover more complete disease trajectories. Second, we relied on one real-world dataset (UTHealth OMOP CDM) to validate the significant disease trajectories. Because this dataset is a single-center data source, data fragmentation may be present. Some patients may have had only intermittent records in the UTHealth OMOP CDM dataset, so diagnoses from admissions at other hospitals may not have been captured. However, even with this limitation, we successfully verified that most significant diagnosis pairs with directionalities for pancreatic cancer (89.9%), STS-TE (98.5%), and STS-AR (83.9%) existed in the respective UTHealth cohorts. Further validation of our method with multi-institutional datasets or other population-based datasets is warranted. Finally, the disease trajectories were made using ICD-10 codes that may not account for more granular clinical signs and symptoms, and our trajectories may not be completely accurate.

### Conclusion

In this study, we proposed a method for discovering signature disease trajectories by leveraging large-scale EHR data. Validation with a local cohort confirmed the feasibility and reliability of the proposed method. Although we demonstrated the validity of our approach with only rare cancers and one real-world dataset, this approach is generalizable to other diseases and other real-world patient datasets. We plan to integrate the knowledge gained from this study to develop EHR-based predictive models to inform early diagnosis, treatment, and prognosis of pancreatic cancer, STS-TE and STS-AR.

## Source

IQVIA Oncology EMR

We thank Madison Semro, Associate Scientific Editor, and Dawn Chalaire, Associate Director of Editing Services, in the Research Medical Library at The University of Texas MD Anderson Cancer Center for editing this article.

## Author Contributions

L.W.: Conceptualized and designed the study, implemented algorithms, analyzed the data, and drafted the manuscript.

R.L.: Designed the study, implemented algorithms, analyzed the data, and drafted the manuscript.

A.W.: Analyzed the data and revised the manuscript.

Q.L.: Revised the manuscript.

J.W.: Revised the manuscript.

X.R.: Revised the manuscript.

C.L.R.: Revised the manuscript.

A.G.: Revised the manuscript.

N.M.: Revised the manuscript.

M.H.G.K.: Revised the manuscript.

Heather Lyu: Designed and supervised the study, analyzed the data, and revised the manuscript.

Hongfang Liu: Supervised the study and revised the manuscript.

## Data Availability

The data that support the findings of this study are available from IQVIA but restrictions apply to the availability of these data, which were used under license for the current study, and so are not publicly available. Data are however available from the authors upon reasonable request and with permission of IQVIA. Other data is provided within the manuscript or supplementary information files.

